# Serum proteomics in COVID-19 patients: Altered coagulation and complement status as a function of IL-6 level

**DOI:** 10.1101/2020.05.29.20116889

**Authors:** Angelo D’Alessandro, Tiffany Thomas, Monika Dzieciatkowska, Ryan C. Hill, Richard O Francis, Krystalyn E. Hudson, James C. Zimring, Eldad A. Hod, Steven L. Spitalnik, Kirk C. Hansen

**Affiliations:** Department of Biochemistry and Molecular Genetics, University of Colorado Denver – Anschutz Medical Campus, Aurora, CO, USA; Department of Pathology & Cell Biology, Columbia University Irving Medical Center, New York, NY, USA; Department of Pathology, University of Virginia, Charlottesville, VA, USA

**Keywords:** SARS-CoV-2, serum, disease severity, clot, inflammation

## Abstract

Over 5 million people around the world have tested positive for the beta coronavirus SARS-CoV-2 as of May 29, 2020, a third of which in the United States alone. These infections are associated with the development of a disease known as COVID-19, which is characterized by several symptoms, including persistent dry cough, shortness of breath, chills, muscle pain, headache, loss of taste or smell, and gastrointestinal distress. COVID-19 has been characterized by elevated mortality (over 100 thousand people have already died in the US alone), mostly due to thromboinflammatory complications that impair lung perfusion and systemic oxygenation in the most severe cases. While the levels of pro-inflammatory cytokines such as interleukin-6 (IL-6) have been associated with the severity of the disease, little is known about the impact of IL-6 levels on the proteome of COVID-19 patients. The present study provides the first proteomics analysis of sera from COVID-19 patients, stratified by circulating levels of IL-6, and correlated to markers of inflammation and renal function. As a function of IL-6 levels, we identified significant dysregulation in serum levels of various coagulation factors, accompanied by increased levels of anti-fibrinolytic components, including several serine protease inhibitors (SERPINs). These were accompanied by up-regulation of the complement cascade and antimicrobial enzymes, especially in subjects with the highest levels of IL-6, which is consistent with an exacerbation of the acute phase response in these subjects. Although our results are observational, they highlight a clear increase in the levels of inhibitory components of the fibrinolytic cascade in severe COVID-19 disease, providing potential clues related to the etiology of coagulopathic complications in COVID-19 and paving the way for potential therapeutic interventions, such as the use of pro-fibrinolytic agents.

## Introduction

In late 2019, a newly identified RNA virus in the family of *Coronaviridae* was identified as the etiology of a form of severe acute respiratory syndrome (SARS).^1^ The 29,903 nucleotides comprising this viral genome share a 89.1% similarity with a group of SARS-like coronaviruses (genus *Betacoronavirus*, subgenus *Sarbecovirus*) that were previously isolated in bats in China.^1^ SARS-CoV-2 viral infection promotes the development of COVID-19 disease, which is characterized by a wide spectrum of clinical symptoms, including fever, persistent dry cough, shortness of breath, chills, muscle pain, headache, loss of taste or smell, and gastrointestinal distress. Although the mechanisms are incompletely understood,^2–4^ the severity of COVID-19 varies across subjects as a function of age and sex (worse in males and older subjects). Various comorbidities worsen the prognosis in COVID-19 patients, including obesity, diabetes, cardiovascular disease, and immunosuppression (e.g., cancer patients undergoing chemo- or radiotherapy or transplant patients).

Structural and protein-protein interaction studies^5^ show that SARS-CoV-2, similar to other beta coronaviruses^6^, enters target cells through the interaction of the viral spike protein S with the host’s angiotensin converting enzyme receptor 2 (ACE2).^7^ ACE2 expression is particularly high in epithelial cells of the oral mucosa^8^ and lung.^9^ Single cell RNAseq studies suggest that heterogeneity in ACE2 expression may contribute to patient-specific and organ-specific responses to this infection.^10^ Blocking the interaction between the ACE2 receptor and spike protein S may be therapeutically useful,^7,11^ as neutralizing antibodies produced in patients tend to recognize epitopes on the spike protein, which is currently the basis for most serological tests that identify previously infected patients and potential convalescent plasma donors.^2,12,13^ Several vaccine candidates are being developed to elicit humoral responses to various capsid proteins, including the spike protein S.^13,14^

Other molecular studies are unraveling the role of other SARS-CoV-2 proteins in mediating the disruption of cellular processes critical to COVID-19 morbidity and mortality. For example, the nucleocapsid (N) protein inhibits type I interferon responses.^15^ Because interferon signaling is critical for the immune response to viral infection,^16^ viral inhibition of this cascade provides a strategy for evading host immune responses. Specifically, the SARS-CoV N protein binds to the SPRY domain of the tripartite motif protein 25 (TRIM25) E3 ubiquitin ligase, thereby interfering with the association between TRIM25 and the retinoic acid-inducible gene I (RIG-I) protein, and inhibiting TRIM25-mediated RIG-I ubiquitination and activation.^15^ Owing to the homology between SARS-CoV-2 and SARS-CoV N proteins, similar mechanisms may help explain the apparent suppression of type I and III interferon responses in COVID-19 patients.^17,18^ Taken together, these considerations support the rationale for therapies aimed at supplementing interferon alpha-2b in these patients.^19^

Beyond viral neutralization strategies aimed at preventing infection, other treatment approaches may decrease viral load and shorten disease duration, such as the antiretroviral drug remdesivir (preliminary results from trial NCT04292899). In addition, other approaches may mitigate the most serious sequelae of SARS-COV-2 infection, which lead to mortality in this population, including inflammatory and coagulopathic complications. For example, severe COVID-19 illness is characterized by the development of “cytokine storm,” characterized by increased circulating levels of pro-inflammatory interleukin-6 (IL-6).^20^ As such, trials are underway to test the efficacy of monoclonal antibodies against the receptor for IL-6 (e.g., tocilizumab) in severe cases of COVID-19.^20,21^ These extreme inflammatory complications in COVID-19 patients are accompanied by lung dysfunction and sustained decreases in oxygen saturation. ultimately resulting in the need for ventilator support or, in extreme cases, extracorporeal membrane oxygenation.^4^ In the most severe cases, ventilation is often (up to ∼80% of the time) insufficient for preventing mortality, owing to the inability to restore lung perfusion resulting from thromboembolic complications.^22^ The observations of a hypercoagulable state^23^ suggested that treatment with anticoagulants (e.g., heparin) or pro-fibrinolytic drugs (e.g., tissue plasminogen activator)^24^ may be beneficial. Although anecdotal evidence about this hypercoagulable state is accumulating, little is known about the molecular factors contributing to this phenotype. Interestingly, platelets from older subjects, in general, are hypercoagulable in the presence of pro-inflammatory stimuli,^25^ which may help explain the increased mortality rates in older COVID-19 patients. Nonetheless, to date, no study has measured coagulation protein levels in COVID-19 patients.

The present study provides the first proteomics analysis of sera from COVID-19 patients, stratified by circulating levels of IL-6, and correlated to markers of inflammation and renal function. As a function of IL-6 levels, we identified significant dysregulation in serum levels of various coagulation factors, accompanied by increased levels of anti-fibrinolytic components, including several serine protease inhibitors (SERPINs). These were accompanied by up-regulation of the complement cascade and antimicrobial enzymes, especially in subjects with the highest levels of IL-6, which is consistent with an exacerbation of the acute phase response in these subjects.^26^ Although our results are observational and preliminary, they clearly demonstrate an increase in the levels of inhibitory components of the fibrinolytic cascade in severe COVID-19 disease. As such, our results support the rationale underlying the potential use of pro-fibrinolytic agents, such as tissue plasminogen activator^24^ or urokinase, in managing coagulopathic complications in severe COVID-19 disease.

## Methods

### Blood collection and processing

This observational study was conducted according to the Declaration of Helsinki, in accordance with good clinical practice guidelines, and approved by the Columbia University Institutional Review Board. Subjects seen at Columbia University Irving Medical Center/New York-Presbyterian Hospital included 33 COVID-19-positive patients, as determined by SARS-CoV-2 nucleic acid testing of nasopharyngeal swabs; in this group, the severity of the disease was inferred from serum levels of IL-6, which were determined by CLIA-certified ELISA-based measurements. The control group included 16 subjects, all of whom were SARS-CoV-2 negative by nasopharyngeal swab at the time of the blood draw. Some patients in this group were “never positive” subjects and some were COVID-19 convalescent patients who were previously positive, but currently negative, and at least 14 days post-resolution of symptoms, as determined by testing nasopharyngeal swabs. Serum was obtained from freshly drawn blood after an overnight hold at 4°C. Sera were then extracted via a modified Folch method (chloroform/methanol/water 8:4:3), which completely inactivates other coronaviruses, such as MERS-CoV^27^. Briefly, 20 μL of serum were diluted in 130 μl of LC-MS grade water, 600 μl of ice-cold chloroform/methanol (2:1) was added, and the samples vortexed for 10 seconds. Samples were then incubated at 4°C for 5 minutes, quickly vortexed (5 seconds), and centrifuged at 14,000 x *g* for 10 minutes at 4°C. The top (i.e., aqueous) and bottom (lipid) phases were removed and the protein disk was further rinsed with methanol (200 ul) prior to centrifugation (14,000 × *g* for 4 minutes) and air drying in a biosafety hood.

### Protein digestion

Protein pellets from serum samples were digested in an S-Trap filter (Protifi, Huntington, NY), following the manufacturer’s procedure. Briefly, ∼50 μg of serum proteins were first mixed with 5% SDS. Samples were reduced with 10 mM dithiothreitol at 55°C for 30 minutes, cooled to room temperature, and then alkylated with 25 mM iodoacetamide in the dark for 30 minutes. Afterward, phosphoric acid was added to the samples to a final concentration of 1.2% followed by 6 volumes of binding buffer (90% methanol; 100 mM triethylammonium bicarbonate (TEAB); pH 7.1). After gentle mixing, the protein solution was loaded onto an S-Trap filter, spun at 2000 g for 1 minute, and the flow-through collected and reloaded onto the filter. This step was repeated three times, and then the filter was washed with 200 μL of binding buffer 3 times. Finally, 1 μg of sequencing-grade trypsin and 150 μL of digestion buffer (50 mM TEAB) were added onto the filter and digested at 47°C for 1 hour. To elute peptides, three step-wise buffers were applied, with 200 μL of each with one more repeat; these included 50 mM TEAB, 0.2% formic acid in water, and 50% acetonitrile and 0.2% formic acid in water. The peptide solutions were pooled, lyophilized, and resuspended in 0.1 % formic acid.

### Nano Ultra-High-Pressure Liquid Chromatography-Tandem Mass Spectrometry (MS) metabolomics

A total of 200 ng of each sample was loaded onto individual Evotips for desalting and then washed with 20 μL 0.1% formic acid followed by the addition of 100 μL of storage solvent (0.1% formic acid) to keep the Evotips wet until analysis. The Evosep One system was coupled to the timsTOF Pro mass spectrometer (Bruker Daltonics, Bremen, Germany). Data were collected over an m/z range of 100 to 1700 for MS and MS/MS on the timsTOF Pro instrument using an accumulation and ramp time of 100 milliseconds. Post processing was performed with PEAKS studio (Version X+, Bioinformatics Solutions Inc., Waterloo, ON). Pathway analyses were performed with the DAVID software and Ingenuity Pathway Analysis. Graphs and statistical analyses were prepared with GraphPad Prism 8.0 (GraphPad Software, Inc, La Jolla, CA), GENE E (Broad Institute, Cambridge, MA, USA), and MetaboAnalyst 4.0.^28^

## Results

### Serum proteomics of COVID-19 patients reveals significant up-regulation of IL-6 targets

An overview of the experimental design is provided in **Figure 1.A**. Briefly, proteomics analyses were performed on sera from 49 subjects, 33 of whom were actively infected with SARS-CoV-2. Results are reported extensively in **Supplementary Table 1**, which identifies 493 proteins (including Uniprot IDs); the Table also includes quantitative values for each protein, as calculated with the PEAKS software by integrating the areas under the curve of all the peptide identifications assigned to a given protein. Volcano plot analyses were performed to highlight proteins that significantly increased or decreased in COVID-19 patient sera (**Figure 1.B**). Pathway analysis of these results confirmed IL-6 signaling as the most upstream up-regulated pathway in COVID-19 patients (*p-value: 3.57 e-17* – **Supplementary Figure 1.A**), as listed in **Supplementary Figure 1.B**. Specifically, several direct and indirect targets of IL-6 signaling were enriched in this dataset, including JNK, STAT3, and p53 (**Supplementary Figure 1.C**). **Supplementary Figure 1.D** provides an overview of the protein targets of IL-6 (and downstream regulators), levels of which increase (red) or decreased (blue) in COVID-19 patient sera.

**Figure 1.**
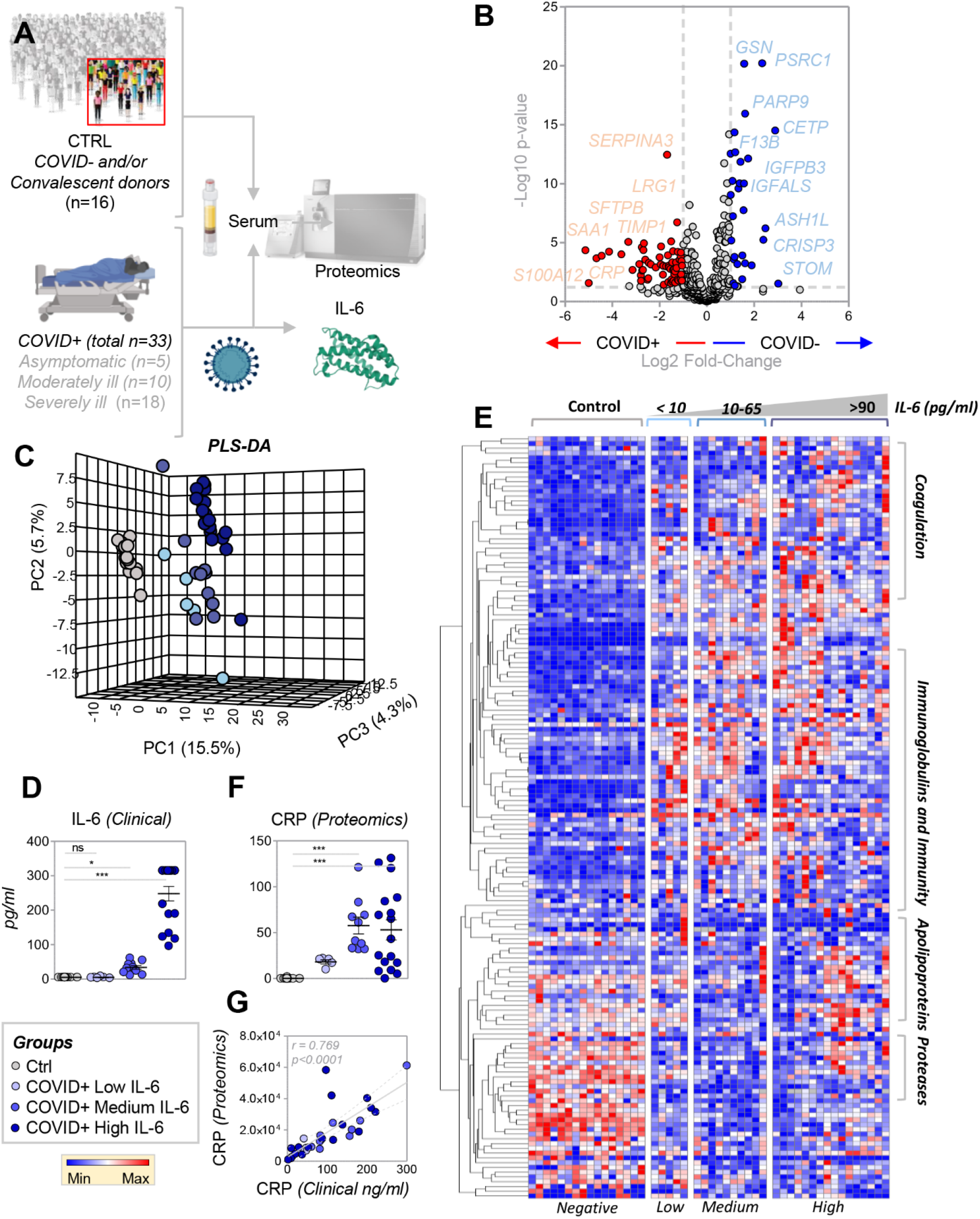
Proteomics analysis of serum from COVID-19 patients. Proteomics analyses were performed on serum from forty-nine subjects, of which 16 were not acutely affected by COVID-19 and 33 were SARS-CoV-2-positive patients, as determined by nucleic acid testing of nasopharyngeal swabs (**A**). Volcano plot analyses in **B** highlight the most significant proteomics changes between the two groups. Following this analysis, we performed a partial least square-discriminant analysis of the data (**C**). As part of this analysis, COVID-19-positive subjects were divided into sub-groups based on IL-6 levels, which were determined during routine clinical care using a clinically-validated ELISA assay (**D**); the patients were classified into groups with low (< 10 pg/ml), medium (10–65 pg/ml), or high (>90 pg/ml) IL-6 levels. Hierarchical clustering based on this classification highlighted a significant impact of COVID-19 and IL-6 levels on proteins involved in inflammation, the complement and coagulation cascades, and antimicrobial enzymes. A vectorial version of this information is provided in **Supplementary Figure 1**. Of note, proteomics results correlated with clinical measurements of the same variable (e.g., creatine kinase M; panel **F**).

### Serum proteomics as a function of IL-6 levels

IL-6 is a marker of COVID-19 disease severity; indeed, clinical trials are underway to disrupt signaling through the IL-6 receptor.^17,18,20,29^ In this context, combined with the pathway analysis described above (**Supplementary Figure 1**), we grouped COVID-19 patients based on IL-6 levels prior to partial least square-discriminant analyses (**Figure 1.C**). Specifically, we used a CLIA-certified clinical assay (**Figure 1.D**) to separate COVID-19 patients into groups with low (< 10 pg/ml), medium (10 < < 65 pg/ml), and high (>90 pg/ml) IL-6 levels. Of note, both PLS-DA (**Figure 1.C**) and hierarchical clustering analyses (**Figure 1.E**) revealed a signature associated with SARS-CoV-2 positivity in the serum proteome (separated across Principal Component 1 (PC1), which explained 15.5% of the total variance; **Figure 1.C**). Consistent with the distribution of samples in the PLS-DA analysis, a subset of proteins with the highest loading weights across PC1 (**Supplementary Table 1**) followed a trend towards progressive increases or decreases as a function of IL-6 levels. One such protein was identified as C-Reactive Protein (CRP; **Figure 1.F**), a marker of the acute phase response for which clinical laboratory values were available and correlated well with proteomics quantification (r = 0.769 *p< 0.0001* – **Figure 1.G**). Pathway analyses, performed on protein clusters in sera of COVID-19 patients classified by IL-6 levels, are shown in a heat map (**Figure 1.E** and **Supplementary Figure 2**); these results indicate a significant enrichment of clusters of proteins related to the coagulation and complement cascades, immunoglobulins and antimicrobial enzymes, apolipoproteins, and other transporters.

### Complement and coagulation cascades

Sera of COVID-19 patients, especially those with IL-6 levels > 10 pg/ml, contained increased levels of multiple proteins in the acute phase response that is initiated by IL-6 – specifically components of the coagulation and complement cascades (top enriched pathway – *p-value: 1.6 e-31* – **Figure 2**). The components that increased or decreased significantly across all groups in this dataset (ANOVA) were annotated in red or blue, respectively, against KEGG pathway hsa04610 (**Figure 2, center panel**; a larger version of this panel is provided in **Supplementary Figure 3**). Specifically, several peptides assigned to coagulation factors (including Factors 2, 5, 7, 10) were increased COVID-19 patient sera, whereas only Factor XIIIb and gelsolin were significantly decreased (**Figure 2**). Interestingly, increases in pro-coagulant components (e.g., kininogen 1 (KNG1), fibrinogen (FGA)) were accompanied by increases in anti-coagulant components (e.g., vitamin K-dependent protein S (PROS1); **Figure 2**). In contrast, sera from COVID-19 patients, especially those with the highest IL-6 levels, exhibited significant increases in serum levels of several SERPINs and carboxypeptidases (CPB2/TAFI) in the coagulation/fibrinolytic^24^ cascade, including SERPINA1, SERPINA3, SERPINF2. Specifically, SERPINA1 – alpha-1 antitrypsin – plays a dual role by slowing down clot formation and inhibiting proteases released by inflammatory cells, like neutrophil elastase. Similarly, SERPINA3 (alpha-1 antichimotrypsin) can inhibit neutrophil cathepsin G and mast cell chymase, both of which can convert angiotensin-1 to the active angiotensin-2.^30^ SERPINF2 (alpha-1 antiplasmin) in an inhibitor of plasmin (and other proteases)^31^ and CPB2 otherwise known as thrombin-activatable fibrinolysis inhibitor (TAFI) both create a pro-thrombotic state by inhibiting the fibrinolytic pathway.

**Figure 2.**
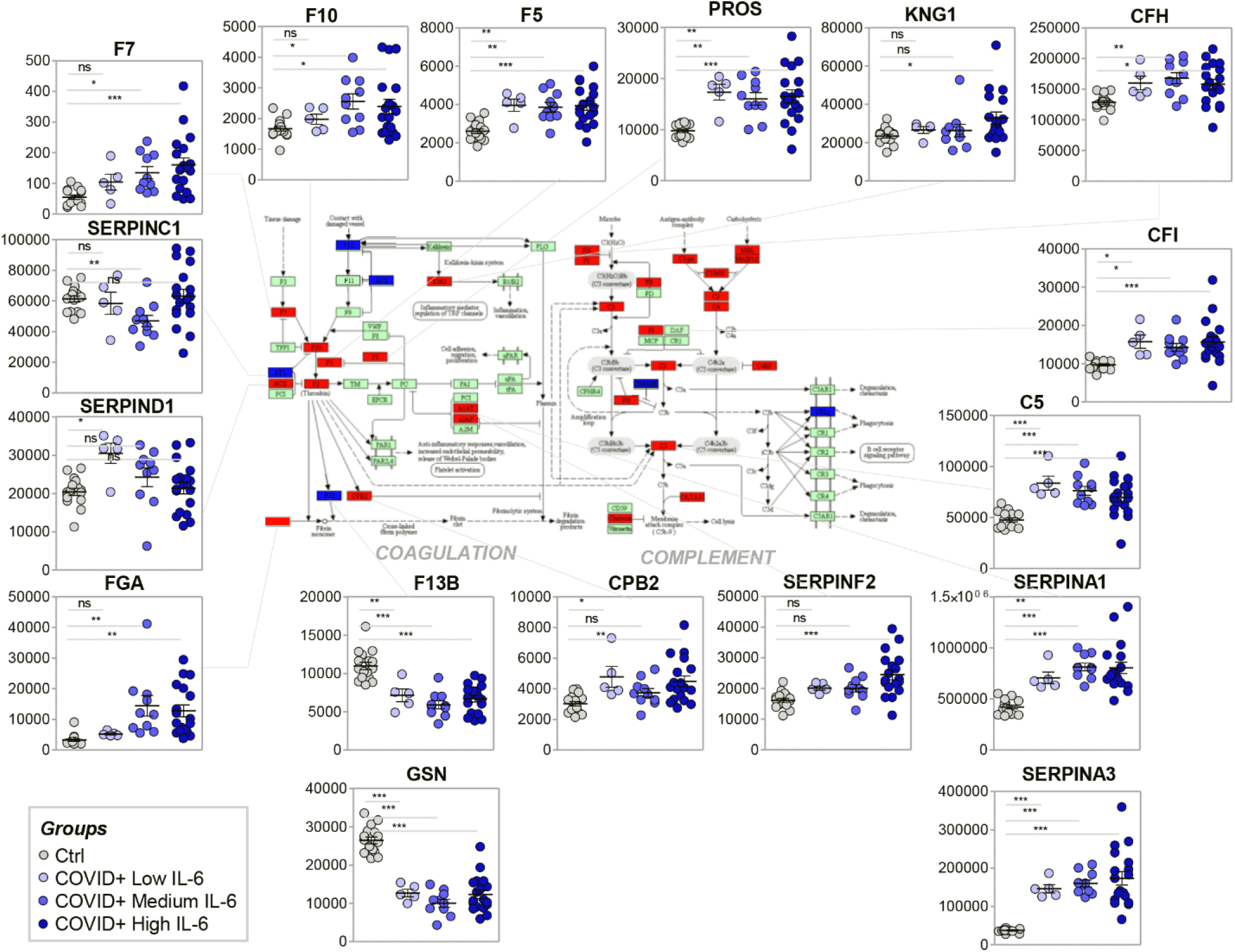
Impact of COVID-19 disease severity (as a function of IL-6 levels) on serum levels of components of the coagulation and complement cascades.

In parallel, several components of the complement cascade were increased in COVID-19 patient sera, including Complement Factor H and I (CFH and CFI, respectively) – that are upregulated to ensure complement targeting of pathogens and not host cells – and C5 (**Figure 2**); this suggests an enhanced innate immune response in these subjects. Of note, deficiency or defects in CFH and CFI are associated with complement-mediated hemolytic uremic syndrome, hemolysis, and thrombosis.^32^

### Immunoglobulins and antimicrobial enzymes

COVID-19 patient sera were significantly enriched in proteins of the adaptive and innate immune responses (**Figure 3**). Although no changes were observed in the levels of immunoglobulin heavy chain constant regions (**Figure 3.A**), all the other components increased. In particular, there were increases heavy chain variable regions (**Figure 3.B**), light chain constant and variable regions (**Figure 3.C-D**), in some cases proportional to IL-6 levels (e.g., IGKV3; **Figure 3.C**). In addition, several enzymes with antimicrobial activity were increased in COVID-19 patient sera, suggesting the possibility of a secondary bacterial infection or simply an exacerbation of the acute phase response (**Figure 3.E**). This trend was particularly evident for cystatin C (CST3), defensin A1 (DEFA1), leucine-rich alpha2 glycoprotein (LRG1), and lysozyme C (LYZC) (**Figure 3.E**).

**Figure 3.**
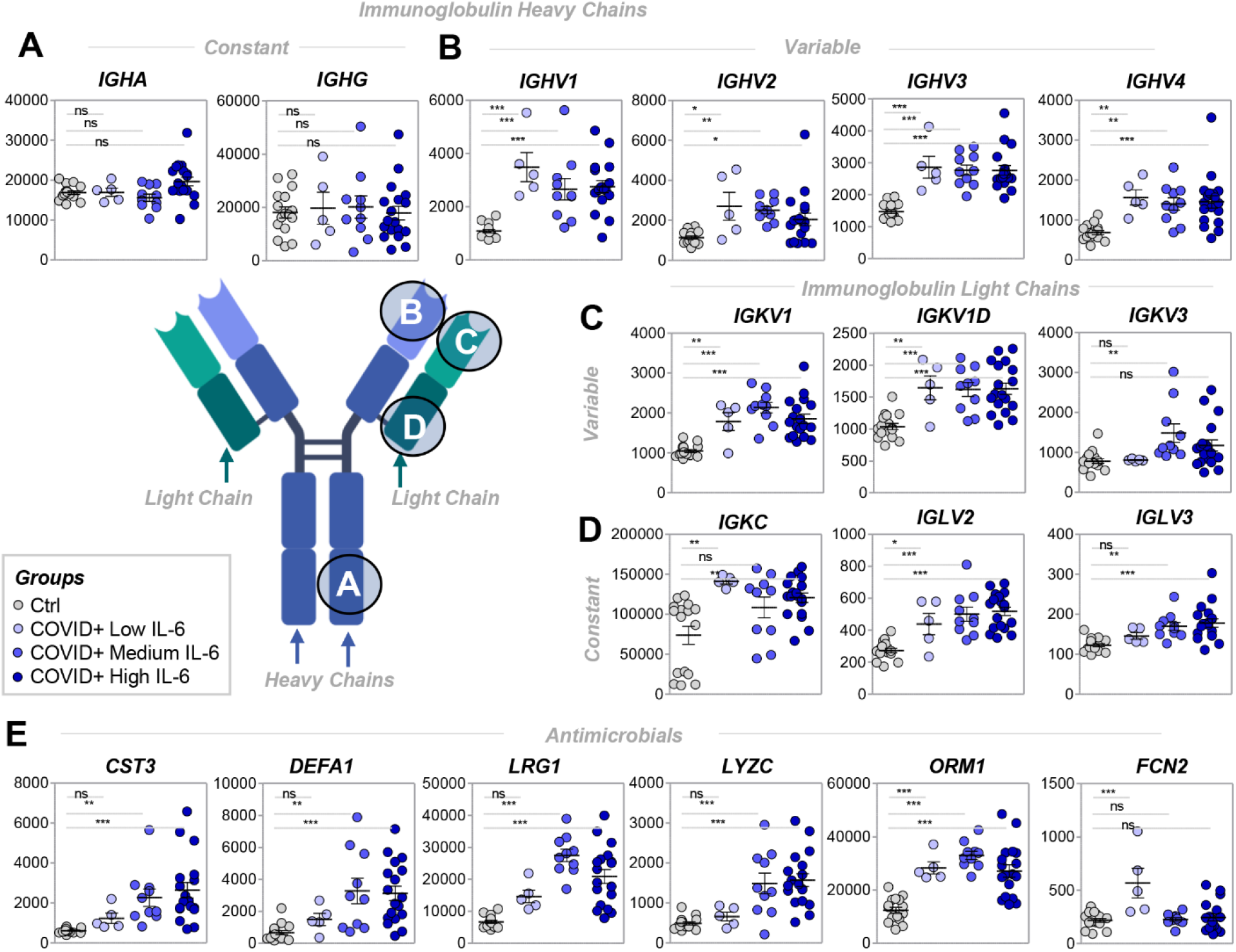
– Impact of COVID-19 disease severity (as a function of IL-6 levels) on serum levels of antibodies and antimicrobial proteins. In COVID-19 patients, significant increases in protein components of immunoglobulin heavy and light chains were identified. Although no significant changes in heavy chain constant regions were observed (**A**), increases were identified in levels of heavy and light variable regions (**BD**) and light chain constant regions (**C**). In **E**, similar increases were observed in levels of antimicrobial enzymes, especially in COVID-19 patients with IL-6 levels > 10 pg/ml (i.e., medium and high IL-6 levels).

### Markers of hemolysis and cell lysis

Sera of COVID-19 patients with the highest IL-6 levels exhibited increased protein markers of hemolysis, including hemoglobin alpha and beta (HBA and HBB) and carbonic anhydrase (CA1; **Figure 4.A**). Other RBC-derived proteins (i.e., band 3 – anion exchanger 1 ((AE1); the most abundant RBC membrane protein), peroxiredoxins 2 and 6, catalase, and biliverdin reductase B) correlated significantly with HBA and HBB levels, despite not reaching significance when compared to COVID-19-negative subjects, suggesting that minimal hemolysis was present in a subset of the most severely ill patients in our study (**Figure 4.A**), perhaps due to mechanical ventilation or other iatrogenic interventions – including the sample collection protocol adopted in this study. In addition, sera from COVID-19 patients with the highest IL-6 levels (>90 pg/ml) had increased levels of the metalloprotease inhibitor TIMP1 and creatine kinase M (CKM; a marker of cardiac tissue damage), but not of actin or histones (**Figure 4.B**).

**Figure 4.**
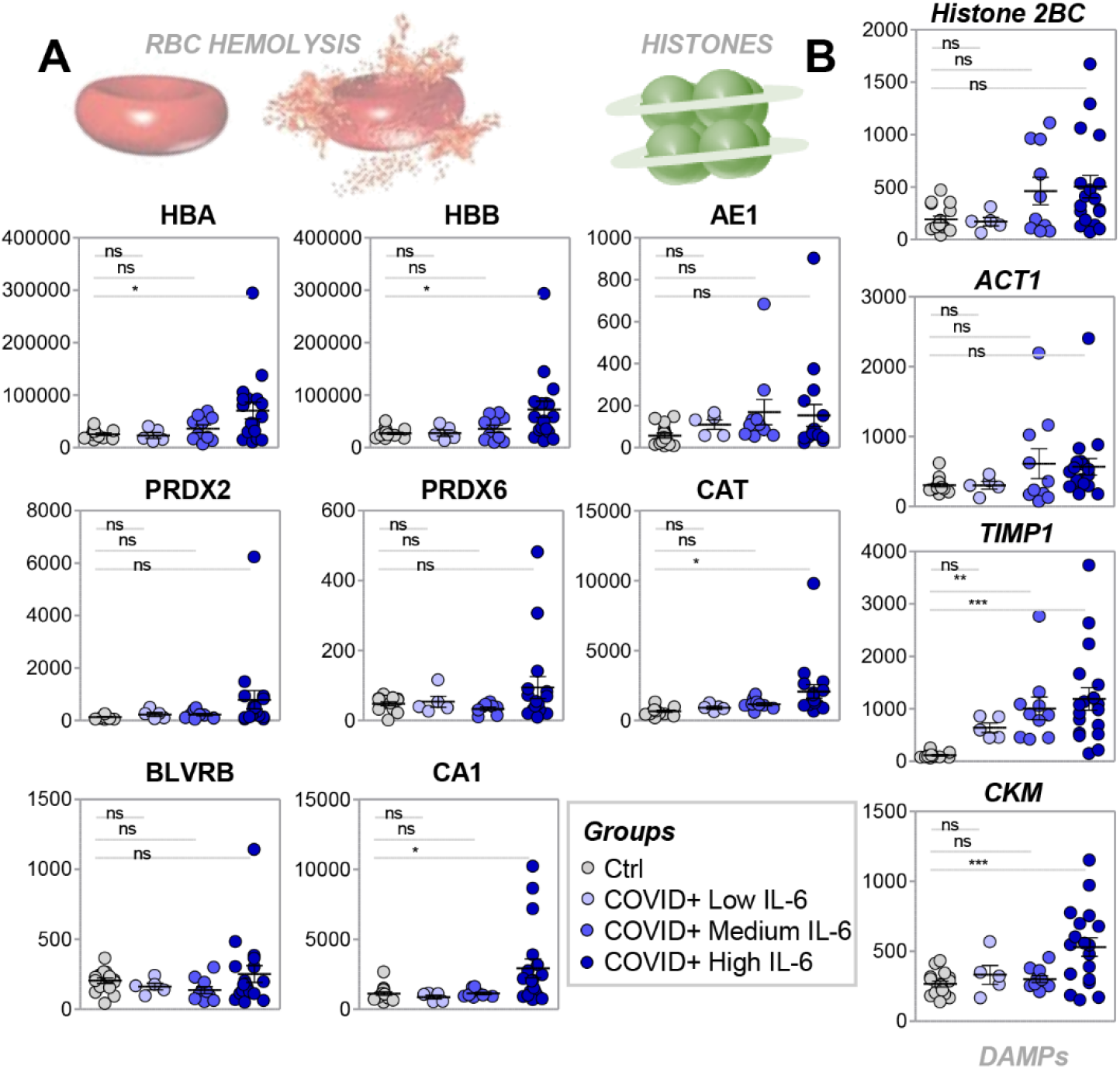
Protein markers of hemolysis (A) or cell lysis (B) increase in the serum of COVID-19 patients with the highest levels of IL-6.

### Proteomic correlates to clinical laboratory parameters of inflammation, cardiac function, and renal function

CKM levels were the top positive correlate to clinical laboratory measurements of IL-6 (**Figure 5.A)**; r = 0.544, *p = 0.001*; **Supplementary Table 1**), followed by proteins involved in extracellular matrix remodeling, such as Inter-alpha-trypsin inhibitor heavy chain H3 (ITIH3), retinol binding protein 4 (RBP4) and complex partner transthyrethin (TTR), collagen 12A1, and glucose phosphate isomerase (GPI; **Figure 5.A**). In contrast, IL-6 levels were negatively correlated to thrombospondin 1 (THBS1; **Figure 5.A**), which has fibrinogen and heparin-binding domains and participates in cell-matrix interactions. Of note, levels of SERPIND1 (heparin cofactor II), a thrombin inhibitor that also binds to, and is activated by, heparin or dermatan sulfate, negatively correlated with IL-6 (**Figure 5.A**). In addition, protein correlates to other inflammatory markers, such as CRP (**Figure 5.B**), showed stronger associations than those observed for IL-6. For example, IL-6, leptin-binding protein (LBP), and complement component C9 all showed strong correlations (>0.75 *p< 0.0001*) with clinical laboratory measurements of CRP levels (**Figure 5.B**; full list in **Supplementary Table 1**), which also correlated well by CRP levels measured by proteomics. Similarly, clinical laboratory measurements of lactate dehydrogenase (LDH; a marker of cell lysis) positively correlated with the levels of LDHA (**Figure 5.C**) and LDHB (**Supplementary Table 1)** as measured by proteomics, catalase (CAT), S100A12, and ezrin (EZR; **Figure 5.C**); the latter is an epithelial cell marker, suggestive of epithelial cell damage, rather than hemolysis, which may also contribute to increased LDH in these patients. In addition, levels of CKM, a marker of cardiac tissue damage, correlated significantly with the proteomics measurements of the same parameter, as well as myoglobin (a marker of myocyte damage; **Figure 5.D**). Finally, creatinine, a marker of renal function, showed significant positive correlations to complement factor D (CFD), a component of the alternative complement activation pathway, as well as LYZC, an antimicrobial enzyme (**Figure 5.E**), suggesting interactions between kidney-related comorbidities and innate immunity in COVID-19 patients.

**Figure 5.**
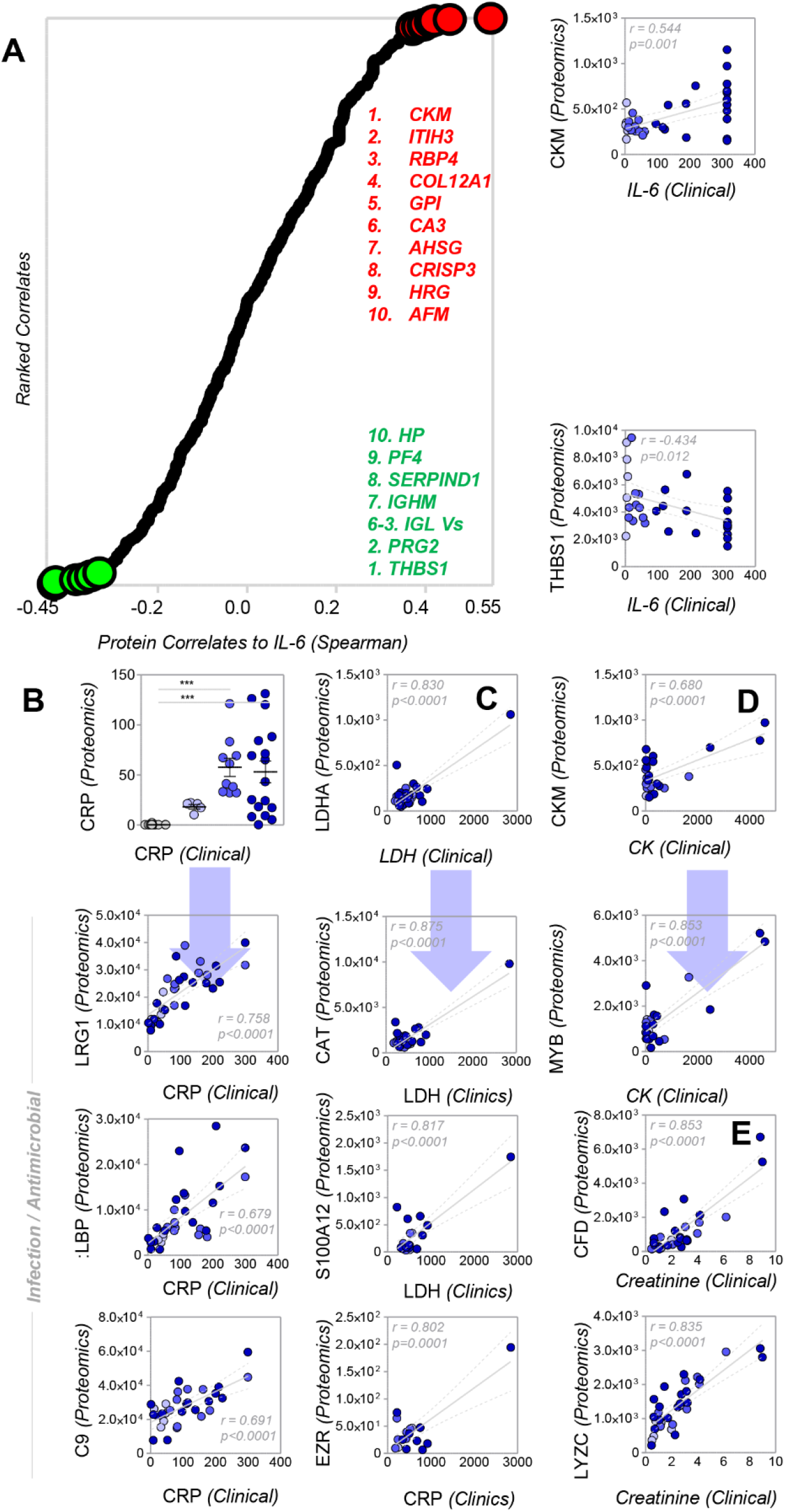
Protein correlates to clinically determined levels of IL-6 (A), CRP (B), lactate dehydrogenase (LDH – C), creatine kinase (D), and creatinine as a marker of renal function (E).

## Discussion

To the best of our knowledge, this work is the first serum proteomics analysis of COVID-19 patients, stratified by the degree of inflammation, represented by IL-6 levels, as a marker of disease severity. Not surprisingly, the degree of inflammation positively correlated with the circulating levels of coagulation components and inhibitors of the fibrinolytic cascade. Although thromboembolic complications have already been reported in the context of COVID-19,^22,24^ the etiology of this phenomenon in this context is unclear. Our results identify increased circulating levels of several coagulation factors, specifically Factor 5, 7, and 10, which could be targeted to prevent untoward clot formation in patients with the highest IL-6 levels. Of note, there is also up-regulation of several SERPIN components in COVID-19 patient sera containing high IL-6 levels. A similar activation of the antifibrinolytic cascade in critically ill trauma patients is associated with the “fibrinolytic shutdown” phenotype.^33^ Thus, this raises the possibility of intervening in COVID-19 patients displaying similar hypercoagulable phenotypes, by restoring the balance between pro- and anti-fibrinolytic cascades by administering pro-fibrinolytic agents, such as tissue plasminogen activator^24^ or urokinase.

Another interesting analogy between COVID-19 patients and trauma patients who develop acute lung injury is the observed increase, proportional to the degree of inflammation, in circulating levels of metalloproteases involved in extracellular matrix remodeling.^34^ Neutrophil^35^ or macrophage^36^ infiltration in the lung secondary to shock, transfusion, hypoxia, and/or inflammation is a hallmark of acute lung injury following trauma/shock, and is a contributor to the early events driving lung fibrosis in pulmonary hypertension^37^ and/or chronic obstructive pulmonary disease.

Our study also identified up-regulation of several components of the complement cascade, along with several enzymes with antimicrobial activity, especially in patients with high IL-6 levels. These observations are directly explained by the role of IL-6 in stimulating acute phase responses– which also has significant crosstalk with the coagulation cascade.^38,39^ Alternatively, these results are suggestive of potential secondary bacterial infection in the severely ill COVID-19 patient which could result in remote organ dysfunction (e.g., kidney), as correlative analysis of proteomics data and creatinine levels seems to suggest. Recent reports from Seattle identified septic shock in a high percentage of ICU patients that required therapeutic intervention to protect the heart and circulatory system.^40^ Of note, IL-6 is also known to stimulate the increase in the levels of several proteases, including matrix metalloproteinases (MMPs), matrilysin (MMP7) and stromelysin-1 (MMP3), which can cleave subclasses of IgG, a consideration that would contribute to explaining the observed increases in the variable chains of light and heavy immunoglobulins, in the absence of increases in the constant fragment of the heavy chain.^41^ Alternatively, coronaviruses have been shown to express papain-like enzymes,^42^ which is relevant in that papains are commonly used in the lab to cleave antibodies into Fab and Fab2’ fragments.

Finally, limited hemolysis was noted in subjects exhibiting the highest levels of IL-6. On the other hand, markers of cell lysis (e.g., LDH) were significantly correlated with markers of epithelial cell damage (e.g., EZR), consistent with the tropism of SARS-CoV-2 for epithelial cells expressing high levels of ACE2 receptor.^7–11,43^ Several of these findings are consistent of the COVID-GRAM scoring system that predicts development of critical illness based on neutrophil-to-lymphocyte ratio, lactate dehydrogenase, and direct bilirubin as 3 of 10 predictive factors.^44^

Nonetheless, there are several limitations that affect the interpretation of the results of the present study. First, the analyses presented herein were performed on residual samples obtained for routine clinical laboratory testing. Serum samples were tested here, which clearly impacts any conclusion related to coagulation cascades. In this view, it is worthwhile to note that any conclusion on coagulation phenotype here drawn based on the measurement of protein levels, not direct determination of enzymatic activity. Although these samples were refrigerated and stored for < 24 hours before initial preparation for analysis, future studies on freshly collected samples will determine whether bias was introduced by these sample collection procedures. In addition, the COVID-19 patients studied herein were mostly male (75%) and older (median age 56), as compared to the controls (62% female, median age 33). Although these sex and age-related biases are consistent with what is observed in clinical cohorts, especially with respect to the most severely ill patients, the relatively limited number of younger, female COVID-19 patients studied here prevented any post-hoc analysis based on age and sex as biological variables; nonetheless, samples are now being prospectively collected to perform these additional studies in newly enrolled cohorts. Moreover, all the COVID-19 patients in this study were inpatients and, as such, they were significantly ill. Future studies will investigate whether asymptomatic, COVID-19-positive patients have proteomic phenotypes comparable to healthy controls or are similar to patients with other, less severe, coronavirus infections. All the patients studied here exhibited symptoms severe enough to require hospital admission; therefore, the relatively small sample size did not allow an analysis focusing on comorbidities and complications (e.g., thromboembolism). To address this issue, longitudinal samples are currently being obtained as part of clinical trials at the Ernest E. Moore Shock Trauma Center at Denver Health (CO, USA) with the goal of investigating the potential for anticoagulants (e.g., heparin) or pro-fibrinolytic agents (e.g., tissue plasminogen activator) to be therapeutically useful in preventing thromboembolic complications in COVID-19 patients.

## Data Availability

All raw data are available as supplementary files

## Acknowledgments

This research was supported by funds from the Boettcher Webb-Waring Investigator Award (ADA), RM1GM131968 (ADA and KCH) from the National Institute of General and Medical Sciences, and R01HL146442 (ADA), R01HL149714 (ADA), R01HL148151 (ADA, SLS), R21HL150032 (ADA), and T32 HL007171 (TN) from the National Heart, Lung, and Blood Institute.

## Authors’ contributions

TT, ROF, SLS, EAH designed the study. TT, ROF, EAH collected and processed the samples. MD, RCH, KCH and ADA performed proteomics analyses and prepared the figures. MD, RCH, KCH and ADA performed data analysis and prepared figures and tables. ADA wrote the first draft of the manuscript, which was significantly revised by SLS, TT, JCZ and KEH and finally approved by all the authors.

